# Distinct Gut Microbiome Signatures in Ethnically Diverse Populations within a Shared Urban Asian Geography

**DOI:** 10.64898/2026.02.06.26345736

**Authors:** Ruwen Zhou, Xiaotao Shen, Theresia H. Mina, Kajal R. Agrawal, Dorrain Y. Low, Jason Xing Kang, Jonathan J. Y. Teo, Shaun H. C. How, Benjamin C. C. Lam, Tong Wang, Chun-Wie Chong, The HELIOS Study Team, Joseph J. Y. Sung, Niranjan Nagarajan, Yusuf Ali, John C. Chambers, Sunny H. Wong

**Affiliations:** Lee Kong Chian School of Medicine, Nanyang Technological University, Singapore; School of Chemistry, Chemical Engineering and Biotechnology, Nanyang Technological University, Singapore, Singapore; Genome Institute of Singapore (GIS), Agency for Science, Technology and Research (A*STAR), Singapore; Integrated Care for Obesity & Diabetes, Khoo Teck Puat Hospital, Singapore; Department of Family Medicine, Changi General Hospital, Singapore; Department of Gastroenterology and Hepatology, Tan Tock Seng Hospital, Singapore; Department of Medicine and Therapeutics, Faculty of Medicine, The Chinese University of Hong Kong; Department of Biological Sciences, Purdue University, United States of America; Monash University Microbiome Research Centre, School of Pharmacy, Monash University Malaysia, Selangor, Malaysia; Department of Biochemistry, Yong Loo Lin School of Medicine, National University of Singapore, Singapore; Clinical Research Unit, Khoo Teck Puat Hospital, National Healthcare Group, Singapore; Precision Health Research (PRECISE), Singapore; Department of Epidemiology and Biostatistics, Imperial College London, United Kingdom

## Abstract

The gut microbiome composition varies across human populations, but its characteristics and determinants in a multi-ethnic setting remain incompletely understood. Singapore, a multicultural city-state, provides a context in which culturally diverse groups share a broadly similar built environment. Using the Health for Life in Singapore (HELIOS) cohort, we profiled the gut microbiome of ethnic Chinese, Indian, and Malay participants (n=861) who resided in the country. Despite substantial overlap in the overall microbial compositions, each group displayed distinct microbial signatures that paralleled culturally rooted dietary habits. Specifically, ethnic Indian participants showed enrichment of multiple *Bifidobacterium* species associated with greater intake of traditional grain-based staples such as idli and thosai; ethnic Malay participants exhibited higher abundance of *Ruminococcaceae* associated with coconut-and rice-based dishes; and ethnic Chinese participants had greater levels of *Bacteroides* associated with seafood- and meat-rich diets. These ethnicity-diet-microbiome relationships were further corroborated by additional data from two independent Malaysian cohorts (n=544), a cohort from the United States (n=210), and an *in vitro* microbial culture model showing selective *Bifidobacterium* expansion by a fermented rice-based batter. Analysis of fecal microbiome-based risk scores revealed ethnic gradients in colorectal neoplasia scores that mirrored population-level cancer incidence patterns. Together, these findings characterize gut microbiome variations across major Asian ethnicities residing in a shared urban environment, providing a reference for precision health strategies relevant to over two billion ethnically relevant people in the Asia-Pacific region and beyond.

## Introduction

The gut microbiome plays a critical role in maintaining health, contributing to essential physiological processes such as digestion, metabolism, and immune function^1,2^. It produces essential nutrients and vitamins, regulates immune responses, and supports the integrity of the intestinal barrier^3,4^. Disruptions in the microbiome, often referred to as dysbiosis, have been implicated in the development of metabolic and digestive disorders, including obesity^5,6^, diabetes mellitus^7^, gastrointestinal cancers^8^, and inflammatory bowel diseases^9^. These insights underscore not only the therapeutic potential of microbiome-targeted interventions, such as dietary strategies and probiotics^10^, but also the urgent need to better understand how the gut microbiome is configured across human populations.

Variations in gut microbiome are known to occur across different populations, influenced by genetic background, lifestyle and environmental factors^11^. Diet is one of the most important determinants, with long-term dietary patterns profoundly influencing microbial composition and functionality^12^. For instance, fiber-rich diets can foster the growth of fermentative bacteria such as *Prevotella* and *Ruminococcus*^13,14^, which effectively metabolize complex carbohydrates into short-chain fatty acids (SCFAs) that help maintain gut barrier integrity, regulate immune responses and provide energy to colonocytes^15,16^. Conversely, diets high in animal fats and proteins are associated with an increase in bile-tolerant bacteria such as *Alistipes* and *Bacteroides*, which may contribute to systemic inflammation and adverse metabolic outcomes^12,17^. Beyond macronutrients, dietary patterns such as a Mediterranean diet, vegetarianism, and the consumption of fermented foods can introduce unique substrates that further modulate gut microbiota, promoting distinct microbial configurations^18,19^.

Gut microbial diversity is closely associated with ethnicity^20^. Distinct ethnic groups harbor unique microbial communities; for example, individuals of African and Asian descent tend to have higher abundances of *Prevotella*, compared to those of European descent where *Bacteroides* is often predominant^20,21^. These differences may arise through the effect of divergent genetic background^22^, dietary variation, environmental factors like sanitation, hygiene practices, and antibiotic use, as well as other exposures that can amplify disparities in gut microbiome composition^23,24^. Lifestyle differences have been shown to drive microbial variations, even in populations within the same provincial region^25^. Additionally, cultural practices, including traditional dietary habits, contribute to microbiome configuration^26^. Such ethnicity-specific microbial signatures directly impact health, potentially influencing susceptibility to diseases such as cardiovascular diseases, digestive cancers, and inflammatory bowel diseases which disproportionately affect certain ethnic groups^27^. However, previous studies on ethnicity-specific differences in gut microbiome have typically involved populations from different regions, where confounding by climate, geography, water supply, and food intake is unavoidable. There remains a critical gap in understanding how diets shape the microbiome in ethnically diverse groups sharing the same environment.

The city-state of Singapore, known for its modern society with a multi-ethnic demographic composition, offers a unique setting of an urban built environment. The three predominant ethnic groups in Singapore, Chinese, Indian, and Malay, exhibit distinct dietary practices and cultural lifestyles, despite residing in proximity within a highly urbanized environment^28^. Chinese diets are typically characterized by a high intake of rice, vegetables, and pork, while Indian diets are rich in spices, legumes, dairy products, and fermented foods such as thosai and idli^29^.In contrast, Malay diets frequently consist of rice, coconut-based dishes, and fish^30^. This juxtaposition of cultural specificity within a shared built environment provides an opportunity to examine how culturally relevant diets relate to gut microbiome, while controlling for geographic and other environmental confounders.

This study aims to explore the relationships between diet, ethnicity, and gut microbiome, using data from multi-ethnic populations within the Health for Life in Singapore (HELIOS) cohort^29^. By providing a large reference of the gut microbiome in Asian populations to date, we addressed a gap in global microbiome science. These data can inform the development of precision nutritional interventions and microbiome-targeted therapies tailored to culturally diverse populations in Asia and beyond.

## Methods

### Study Design and Participant Characterization

This study included 861 participants from the Health for Life in Singapore (HELIOS) cohort (ethics approval IRB-2016-11-030), a population-based longitudinal study designed to identify environmental, lifestyle, and genetic factors contributing to chronic diseases in Singapore’s multi-ethnic population. The cohort recruited more than 50,000 Singapore citizens or permanent residents across diverse socioeconomic backgrounds, aged 30–84 years old. Participants were recruited from the general population and excluded at enrollment if they were pregnant or breastfeeding, had a history of major illness requiring hospitalization or surgery, had received cancer treatment within the past year, or had recently participated in clinical drug trials^29^. For the current analysis, participants of Chinese (n = 683), Indian (n = 109), and Malay (n = 69) ethnicity were included based on the availability of dietary, stool microbiome, and clinical data. Comprehensive demographic data was collected, including age, sex, and self-reported ethnicity. Trained personnel performed standardized anthropometric measurements including height, weight, waist circumference, and hip circumference. Body mass index (BMI) was calculated as weight in kilograms divided by height in meters squared (kg/m^2^). Fasting blood samples were collected from all participants.

Dietary intake was assessed using a locally validated 253-item Food Frequency Questionnaire (FFQ) developed for Singapore’s multi-ethnic population^30^. Participants were asked to recall their habitual food consumption, reporting both portion sizes and frequency of consumption for each item. Nutrient values were calculated from FFQ, using Singapore Health Promotion Board composition database (https://www.hpb.gov.sg/healthy-living/food-beverage/tools), to estimate intakes of 26 macronutrients (*e.g.*, carbohydrates, protein, total fat, fiber) and micronutrients (*e.g.*, vitamins and minerals).

### Metagenomic Sequencing and Microbial Profile

Stool samples were self-collected by participants using the Fe-Col® collector (Alpha Laboratories) and DNA/RNA Shield™ Fecal Collection Tubes (Zymo Research). Approximately 1 g of fecal material was transferred into tubes prefilled with DNA/RNA Shield™ to inactivate microorganisms and preserve nucleic acids at ambient temperature, followed by thorough mixing. Samples collected at home were returned by mail and subsequently stored at −80 °C until DNA extraction. Genomic DNA was extracted from stool samples using the QIAamp PowerFecal Pro DNA Kit (Qiagen) following the manufacturer’s instructions. Metagenomic libraries were prepared using the NEBNext Ultra II FS DNA Library Prep Kit (New England Biolabs) and sequenced on an Illumina HiSeq X platform to generate 2×150 bp paired-end reads, yielding an average of 22 million reads per sample. Raw paired-end FASTQ reads from shotgun metagenomic sequencing were quality-filtered using fastp^31^ v0.20.1. Quality control steps included adapter trimming, removal of low-quality bases (Phred score <20), and exclusion of reads shorter than 50 base pairs. To remove host contamination, filtered reads were aligned to the human reference genome (hg19/GRCh37) using BWA-MEM v0.7.17^32^, and unmapped reads were retained as putative microbial sequences. Taxonomic classification of microbial reads was performed using Kraken2^33^ v2.0.8 with the standard 16 GB database (release date: 2022-12-09), which contains reference genomes from bacteria, archaea, viruses, and selected eukaryotic organisms^33^. Species- and genus-level relative abundance estimates were refined using Bracken v2.5 (Bayesian Reestimation of Abundance with KrakEN)^34^, which uses a Bayesian approach to redistribute ambiguous k-mer assignments and improve taxonomic resolution.

Functional gene profiling with the metagenomic data was conducted using HUMAnN3^35^ (HMP Unified Metabolic Analysis Network) v3.1.0 in two stages. First, reads were aligned to the ChocoPhlAn^35^ pangenome database (species-level pangenomes) using nucleotide-based search to identify known microbial genes; subsequently, unmapped reads underwent translated search (six-frame translation) against the UniRef90^36^ protein database to capture novel or divergent sequences. Gene family abundances were quantified and normalized to copies per million (CPM) to account for differences in sequencing depth across samples.

### Microbiome Diversity and Composition Analyses

Gut microbiome composition was characterized using alpha- and beta-diversity metrics. Alpha diversity, which quantifies within-sample microbial diversity, was evaluated using multiple complementary indices: the Chao1 index for species richness^37^; the Shannon^38^ and Gini-Simpson^39^ indices for both richness and evenness, with Shannon emphasizing rare species and Gini-Simpson weighting dominant species; and the Fisher index^40^ for modeling the species abundance distribution. Statistical comparisons were performed using pairwise Wilcoxon rank-sum tests, with *p*-values adjusted for multiple testing using False Discovery Rate (FDR) correction at (statistical significance was defined as *q* < 0.05). Beta diversity was visualized using Principal Coordinate Analysis (PCoA) based on Bray-Curtis dissimilarity matrices^41^, which quantifies compositional differences between samples. Statistical significance of community-level differences was assessed using Permutational Multivariate Analysis of Variance (PERMANOVA)^42^ analysis.

To identify differentially abundant microbial species across clinical or ethnic groups, we employed the DESeq2 v1.34.0 framework^43^, which models count data using negative binomial generalized linear models with shrinkage estimation for dispersions and fold changes. Species with FDR-adjusted *q*-values < 0.05 were considered significant. Diet-microbiome associations stratified by ethnicity were visualized using volcano plots, highlighting species with adjusted *q* < 0.05 and absolute fold change ≥ 2 as significantly associated.

### Dietary Data and Microbiome Association Analyses

Food intake was analyzed using the FFQ data, covering commonly consumed food groups including bakery bread, idli/thosai, lontong, root vegetables, coffee, tea, dairy, legumes, chapati, fruits, rice dishes, mushrooms, pork, red meat, seafood, poultry, leafy vegetables, cruciferous vegetables, noodle dishes, and eggs in the unit of serve per day. They were selected based on both ethnically differential consumption patterns and the most commonly consumed foods across the cohort. Differences in food group intake across ethnic groups were assessed using the Kruskal–Wallis test for global comparisons, followed by pairwise Wilcoxon rank-sum tests for post hoc analysis. All *p*-values were adjusted for multiple comparisons using the Benjamini–Hochberg (BH) correction.

To investigate associations between dietary intake and microbial species abundance, we performed Spearman rank correlation analysis adjusted for potential confounders (age, sex, BMI, and ethnicity) using partial correlation. To reduce sparsity bias and improve statistical robustness, before all downstream analysis, microbial species with ≥10% prevalence across samples and a minimum relative abundance of 0.001% were included in the analysis.^44,45^. Results were visualized using heatmaps, with significant associations defined at FDR-adjusted *q <* 0.05 and color-coded by correlation direction.

To visualize ethnic-specific patterns in dietary intake and gut microbiome composition, ternary diagrams were constructed based on the relative contributions of the three major ethnic groups. For dietary analysis, per-capita intake values were calculated for each food item across individuals within each ethnic group. These values were then normalized such that, for each food item, the summed contribution across the three ethnic groups equaled 100%. Each food item was represented as a point within the ternary space, positioned according to its relative proportional intake among Chinese, Indian, and Malay participants. Food items were colored according to predefined food categories (e.g., carbohydrates, plant-based foods, protein, dairy, beverages) to facilitate interpretation of dietary pattern clustering. Similarly, for microbiome analysis, per-capita relative abundances of microbial species were calculated within each ethnic group and normalized across the three groups to obtain proportional contributions summing to 100% per species. Microbial points were colored by bacterial taxonomic class to highlight compositional differences at higher taxonomic levels. For both analyses, points located closer to a given vertex indicate relative enrichment in the corresponding ethnic group, whereas points near the center represent features shared more evenly across groups.

Correlation networks were constructed to visualize the relationships between food intake, nutritional factors, microbial species, and ethnicity. Network edges represented statistically significant correlations (adjusted *q <* 0.05), with edge color indicating correlation direction (red for positive, blue for negative) and edge thickness proportional to correlation strength. Each node represents a food item, nutrient, microbial species, or ethnic group, with node size reflecting its degree (number of connections).

### Microbiome-Based Clinical Risk Scoring

To evaluate the potential health implications of gut microbiome variations in our cohort, we assessed disease risk profiles using established microbiome-based signatures for five gastrointestinal disorders: colorectal cancer (CRC), colorectal adenoma, Crohn’s disease, ulcerative colitis, and irritable bowel syndrome. We computed microbial risk scores derived from published metagenomic case–control studies, based on a 50-species multinomial LASSO classifier which provides species-specific coefficient weights^46^ for multiple diseases. Another score was derived from a 29-species CRC signature identified through a large multi-cohort meta-analysis^47^. For both models, individual risk scores were computed by multiplying the relative abundance of each signature species by its corresponding published coefficient and summing the weighted values to obtain a composite index for each participant. These microbiome-derived scores were then compared by Wilcoxon rank-sum test across ethnic groups to assess potential ethnicity-specific differences in microbiome-associated disease risk profiles.

### Validation Cohorts

To validate our findings on gut microbiome associations across Chinese, Indian, and Malay ethnic groups, we utilized 16S rRNA amplicon sequencing data from two Malaysian cohort studies. The first dataset comprised 214 community-dwelling individuals, including 65 Chinese, 46 Malay, and 49 Indian individuals^26^. The second dataset was obtained from the 1000MY Microbiome Study (BioProject: PRJNA1179675), which included 238 Chinese, 94 Malay, and 52 Indian individuals^48^. After combining the two datasets, a total of 544 samples from ethnic Chinese, Malay, and Indian participants were included in the validation analysis.

Raw sequence data from both studies were processed using a standardized analytical pipeline to ensure comparability. Quality-filtered reads were denoised using DADA2^49^ to generate amplicon sequence variants (ASVs), followed by taxonomic classification using the Greengenes2^50^ reference database, and agglomeration at the genus level for consistency with available reference annotations. To account for compositional constraints and sequencing depth variation, we applied centered log-ratio (CLR) transformation after zero replacement using the geometric Bayesian multiplicative replacement method from the R package zCompositions. Technical batch effects between cohorts were corrected using ComBat from the sva R package prior to downstream analyses.

We evaluated the reproducibility of ethnicity-associated microbial patterns by comparing diversity metrics and taxonomic profiles between the validation cohorts and our primary study population. Specifically, we examined the relative abundance of *Bifidobacterium* and other identified taxa to assess the reproducibility of ethnicity-associated microbial signatures across independent, multi-ethnic Southeast Asian populations. Statistical comparisons were performed using the same methods as the primary analysis to ensure methodological consistency.

To validate the association between *Bifidobacterium* abundance and traditional Indian grain-based staple foods observed in the HELIOS cohort, we analyzed a separate longitudinal dataset from the United States^51^. This study included individuals with time-series measurements, consisting of paired daily dietary recalls collected via the Automated Self-Administered 24-Hour Dietary Assessment Tool^52^ (ASA24) and gut microbial compositions profiled with 16S rRNA sequencing. To account for dietary transit time, each dietary intake record from day *t* was paired with the gut microbial composition from day *t+1*. For this validation, subjects were categorized as chapati consumers or non-consumers based on their reported intake. Differences in *Bifidobacterium* abundance between these groups were assessed using the Wilcoxon rank-sum test, and distributions were visualized using boxplots showing median and interquartile ranges.

### In Vitro Microbial Culture Experiment

Thosai, a traditional Indian fermented rice-based batter, was obtained from a local commercial establishment in Singapore to represent a standard consumer serving. Typical preparation of this staple involves a batter of wet-ground rice (*Oryza sativa*) and decorticated black gram lentils (*Vigna mungo*), mixed with fenugreek seeds and salt, fermented overnight at ambient temperature, and cooked on a hot griddle. Digestion of thosai was performed following the standardized INFOGEST 2.0 static *in vitro* digestion protocol to simulate sequential oral, gastric, and intestinal phases^53^. Briefly, homogenized thosai samples were mixed with simulated salivary fluid containing α-amylase and incubated at 37 °C for 2 min (pH 7.0) to initiate oral digestion. Gastric digestion was conducted by adjusting the pH to 3.0, adding pepsin (3,000 U/mL), and incubating for 2 hours under constant agitation at 37 °C. The intestinal phase was initiated by neutralizing the pH to 7.0 and supplementing it with pancreatin and bile salts, followed by a further 2-hour incubation at 37 °C to simulate small-intestinal conditions. At the end of digestion, enzyme activity was terminated by adding trichloroacetic acid (TCA), and the digesta were subsequently centrifuged and freeze-dried to obtain a powdered form. The dried digesta were stored at −80 °C until further use.

To examine the impact of digested thosai on gut microbial dynamics, we employed a stool-derived *in vitro* community (SIC) microbial culture model^54^. Fresh fecal samples from four healthy human donors (aged 25–35 years with normal BMI and no antibiotic use within 6 months), each confirmed to contain *Bifidobacterium* species from metagenomic sequencing, were used as inocula. It is approved by Nanyang Technological University (IRB-2023-747). The SIC cultures were maintained under strict anaerobic conditions at 37°C in Bioreactor Medium (BRM). To sustain microbial activity and community stability, cultures were serially passaged every 48 hours by 1:100 dilution into fresh medium. The system was propagated for nine passages (P0–P9). At passage 5 (P5), thosai digesta was introduced into the cultures at a final concentration of 1% (v/w) as the treatment condition, while control cultures received BRM alone without digesta supplementation.

Samples were collected at passages P3, P5, P7, and P9 for microbial profiling. Genomic DNA was extracted using a commercial fecal DNA extraction kit (QIAamp PowerFecal Pro DNA Kit, Qiagen) according to the manufacturer’s instructions. Shotgun metagenomic sequencing was performed on an Illumina NovaSeq 6000 platform (2 × 150 bp paired end reads) to achieve comprehensive taxonomic and functional characterization of the microbial communities. Raw sequencing reads were subjected to quality control using fastp^31^ (v0.20.1) to trim adapters and filter low-quality bases. Taxonomic classification was performed using Kraken2^33^. Functional gene profiling was subsequently conducted using HUMAnN3^35^ for taxonomic stratification and the UniRef^55^ database for gene family mapping. Species-level relative abundances were derived to evaluate the growth response and compositional shifts of *Bifidobacterium* spp. across donors, treatments, and passages. Statistical assessment of *Bifidobacterium* dynamics across SIC passages was performed using Wilcoxon rank-sum tests for pairwise passage comparisons, complemented by Spearman correlation to evaluate monotonic trends and linear regression models adjusting for donor effects.

## Results

### Participant Characteristics

A total of 861 participants were included in the primary analysis. The mean age was 50.0 ± 11.4 years, and 62.6% of participants were female. The cohort comprised primarily of ethnic Chinese individuals (n=683, 79.3%), followed by ethnic Indian (n=109, 12.7%) and Malay (n=69, 8.0%) participants (**Fig. 1**, **Table 1, Supplementary Table 1**). The mean body mass index (BMI) was 24.9 ± 4.7 kg/m^2^. The cohort included participants from all three major ethnic groups in Singapore, enabling comparative analyses across ethnicities.

**Figure 1.**
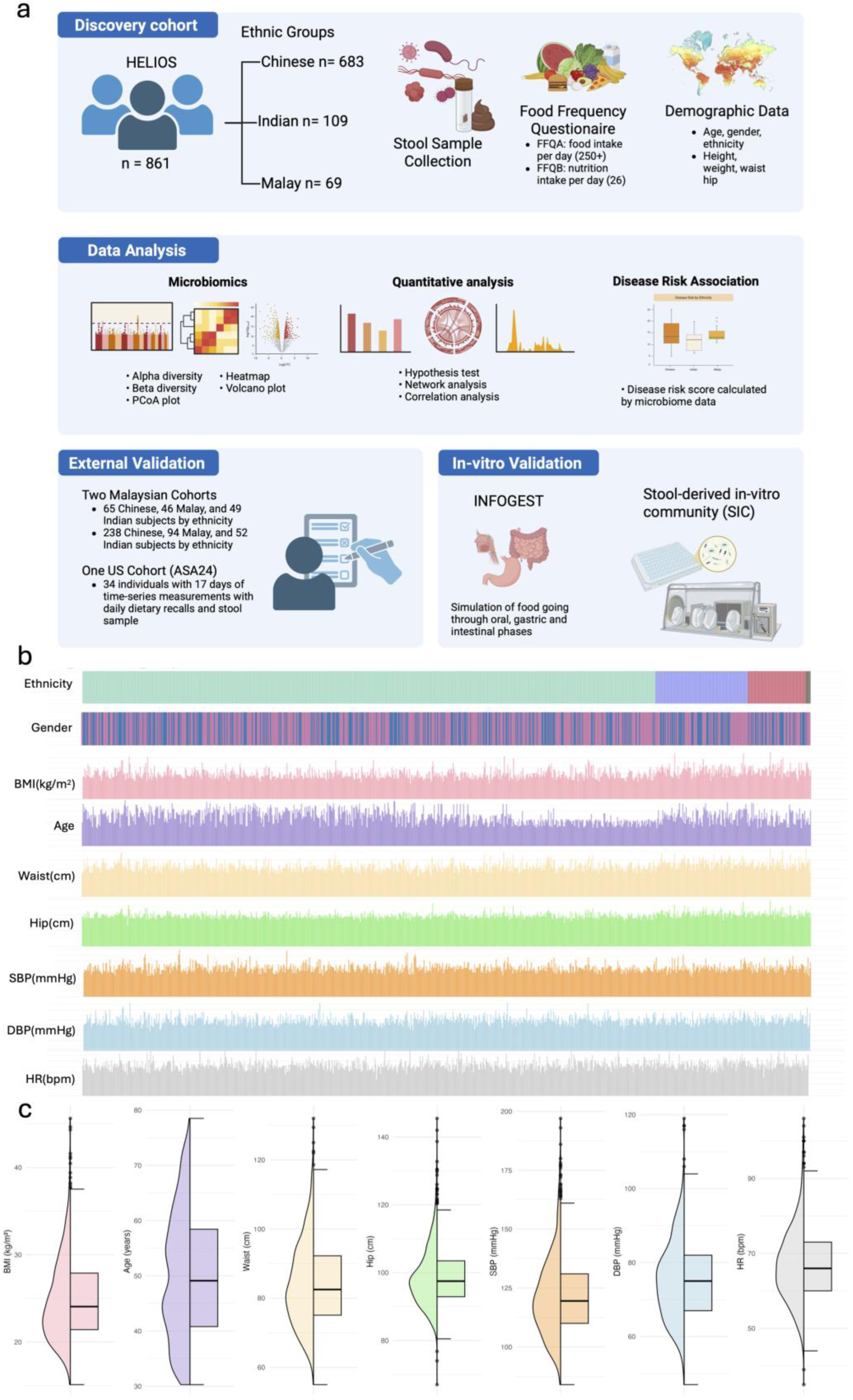
Study workflow and demographic characteristics of participants. (a) The workflow spans participant recruitment from the HELIOS discovery cohort and subsequent analysis. Findings were validated using two external datasets from Malaysia^26,48^ and one from the US (ASA24)^51^, followed by experimental verification using the INFOGEST in-vitro digestion model^53^ and a Stool-derived In-vitro Community (SIC)^54^. (b) Demographic and clinical characteristics of the 861 participants, stratified by ethnicity. Chinese with green, Indian with purple and Malay with red. Key variables include ethnicity, gender, age, body mass index (BMI), hip circumference, waist circumference, systolic blood pressure (SBP), diastolic blood pressure (DBP), and heart rate (HR). (c) Violin plots depicting the distribution of key anthropometric and clinical parameters across the study population

**Table 1:**
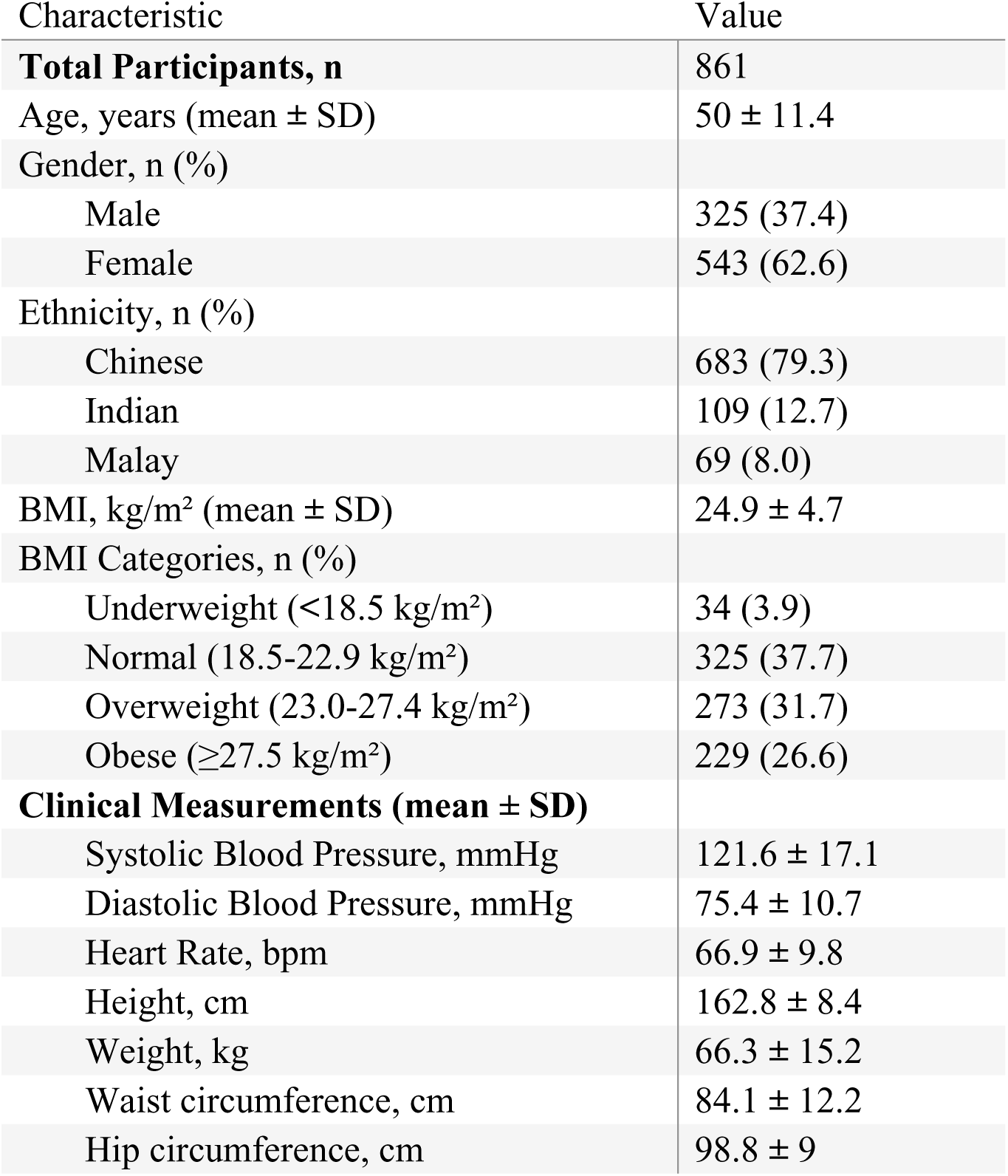
Characteristics table of study participants (n=861). Data are presented as mean ± standard deviation (SD) for continuous variables and n (%) for categorical variables. BMI categories follow WHO Asian-specific cutoffs.

### Gut Microbial Diversity Across Ethnic Groups

Microbial richness, as measured by the Chao1 index, did not differ significantly across ethnic groups (**Fig. 2a**). However, the Shannon and Gini-Simpson indices were significantly higher in Malays compared to Indian participants (both *p <* 0.05), indicating greater microbial diversity (**Fig. 2a**). Despite consideration overlap in the PCoA clustering pattern between ethnic groups, the Bray–Curtis dissimilarity demonstrated modest but significant separation between ethnic groups, with significant differences in overall composition (PERMANOVA *p <* 0.001) (**Fig. 2b**). While ethnicity contributes to microbiome structure, inter-individual variability within ethnic groups remains substantial. The mean Bray–Curtis dissimilarity within ethnic groups (0.721 ± 0.124) was comparable to that observed between ethnic groups (0.753 ± 0.124), further emphasizing the high degree of interpersonal heterogeneity in microbiome composition within this shared urban setting.

**Figure 2.**
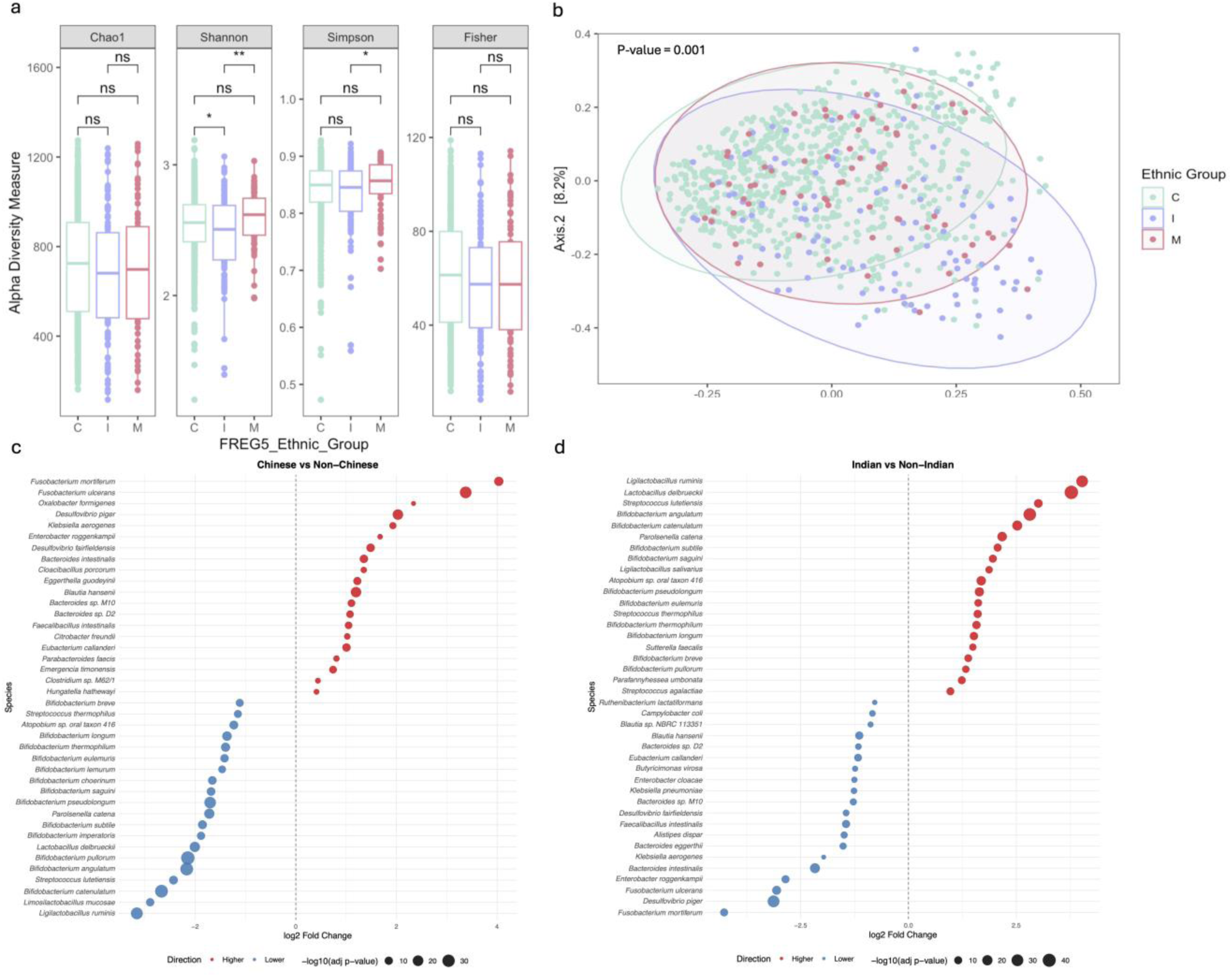
Ecological diversity and microbiome composition across ethnic groups. (a) Alpha diversity metrics, including Chao1, Shannon, Gini-Simpson, and Fisher indices, comparing microbial diversity among the three ethnic groups. (b) PCoA illustrating differences in microbiome composition among the three groups, with statistical comparisons performed using PERMANOVA. (c) Differential abundance analysis of microbiome taxa comparing Chinese and non-Chinese individuals, visualized as a lollipop plot. Each point represents a microbial species, with the x-axis indicating the effect size (log₂ fold change). Point size corresponds to the mean relative abundance of each species in the dataset. Species are ordered vertically according to effect size, with labels provided for the most strongly enriched taxa. Positive (red) log₂ fold-change values represent higher abundance in Malays, whereas negative (blue) values represent enrichment in non-Malays. (d) Differential abundance analysis of microbiome taxa comparing Indian and non-Indian individuals, visualized as a lollipop plot.

### Ethnicity-Specific Microbial Signatures

Despite the broad overlap in clustering patterns, differential abundance analysis identified clear ethnicity-specific microbial signatures (**Figs. 2c-d, Supplementary Figs. 1-2, Supplementary Table 2)**. Indian participants were enriched for multiple *Bifidobacterium* species, such as *Bifidobacterium subtile* (log of fold change (logFC)=1.38, *q*=0.002)*, Bifidobacterium saguini* (logFC=1.31, *q*=0.001), *Bifidobacterium pseudolongum* (logFC = 1.22, *q* = 0.004), and *Bifidobacterium longum* (logFC=1.06, *q*=0.02). Chinese participants showed reduction of *Ligilactobacillus ruminis* (logFC = -2.87, *q*=0.03), and *Bifidobacterium catenulatum* (logFC =-2.24, *q* =0.01), alongside enrichment in several *Bacteroides* and *Fusobacterium* species. These patterns indicate that the ethnic groups retain distinctive microbial features that reflect underlying cultural and dietary practices.

To test the reproducibility of these findings, we analyzed an independent validation dataset comprising 544 participants from two Malaysian population cohorts, which included a comparable composition of ethnic Chinese, Indian, and Malay individuals. Consistent with our primary analysis, Indian participants exhibited lower microbial richness (Chao1 and Fisher indices) and diversity (Shannon and Simpon indices) (all *p <* 0.001). Beta diversity analysis confirmed significant compositional differences among ethnic groups (PERMANOVA *p <* 0.001), with PCoA plots showing similar clustering patterns across ethnicities (**Supplementary Fig. 3**). At the taxonomic level, we observed higher abundance of *Bifidobacterium* among Indian participants compared with Chinese and Malay participants (*q <* 0.01, **Supplementary Fig. 3a**).

### Diet-Microbiome Associations

We explored the associations between specific food groups and the abundance of ethnicity-associated bacterial taxa, using Spearman correlation adjusted for age, gender, ethnicity, and BMI. Multiple *Bifidobacterium* species showed positive associations with consumption of bakery bread and grain-based staple foods, which remained robust after multiple testing correction (**Fig. 3a, Supplementary Table 3**). These include associations with bakery bread (*B. catenulatum*: *r*=0.161, *q=*2×10^-6^; *B. angulatum*: *r*=0.139, *q=*4.8×10^-5^), Indian staples such as idli and thosai (*B. angulatum*: *r* = 0.133, *q =* 9.5×10^-5^), chapati (*B. catenulatum*: *r*=0.07*, q*=3×10^-2^), and a common Malay food of lontong, which is rice cake (*B. adolescentis*: *r*=0.125, *q=*2.5×10^-4^). These observations hinted a potential link between common grain-based staple foods and *Bifidobacterium* in our digestive tract^56,57^, and contextualized the culturally rooted dietary practices with the gut microbial profiles.

**Figure 3.**
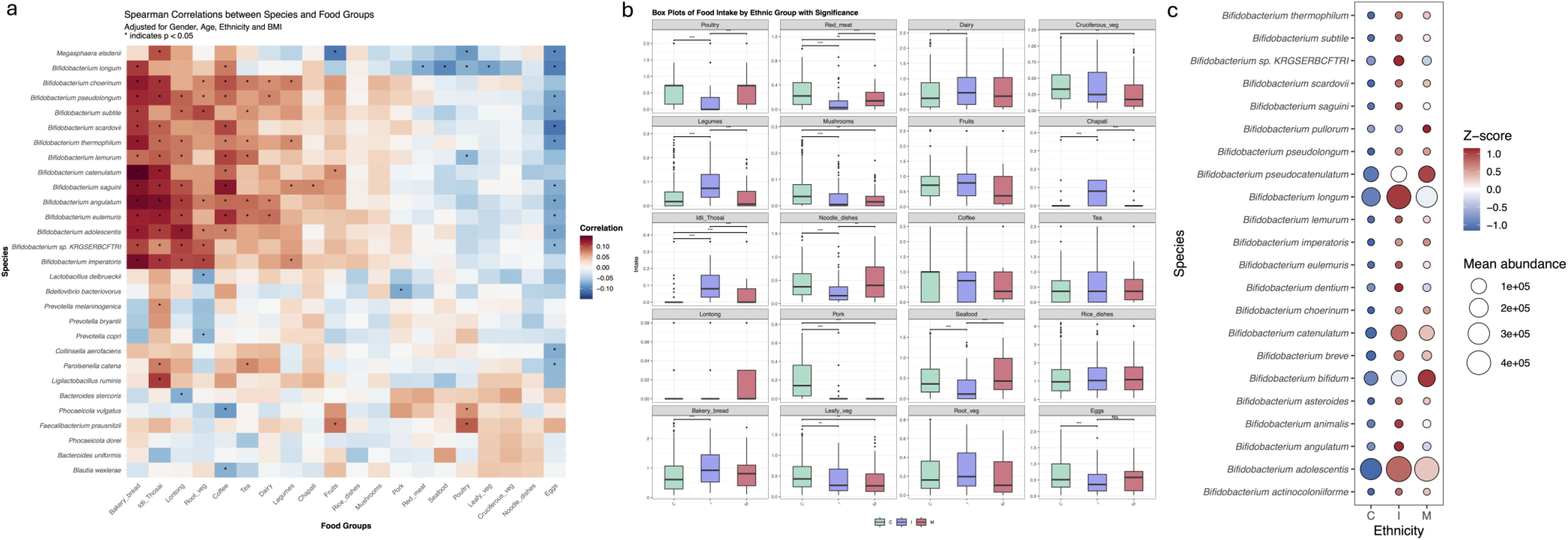
Correlation between ethnicity-associated microbiota and dietary intake. (a) Spearman correlation heatmap depicting the associations between the relative abundance of ethnicity-specific bacterial taxa and daily intake across various food categories, adjusted for age, gender, ethnicity, and BMI. Significant correlations are indicated by asterisks (*, FDR q<0.05). Hierarchical clustering of both bacterial taxa and dietary variables reveals patterns of co-occurrence. Species selected based on both statistical significance of differential abundance across ethnic groups and top ten most abundant species within the HELIOS cohort, including *Faecalibacterium prausnitzii, Phocaeicola vulgatus, Prevotella copri, Bacteroides uniformis, Bifidobacterium adolescentis, Phocaeicola dorei, Blautia wexlerae, Bifidobacterium longum, Collinsella aerofaciens, Bacteroides stercoris*. (b) Box plots illustrating the distribution of dietary intake across the three ethnic groups (Chinese [C], Indian [I], Malay [M]). Statistically significant differences were assessed using the Kruskal-Wallis test, with significance levels denoted as * (p<0.05), ** (p<0.01), and *** (p<0.001). (c) Bubble heatmap showing the abundance of *Bifidobacterium* species across ethnic groups. Circle size reflects the raw mean abundance of that species within the ethnic group. Circle color denotes the standardized abundance (z-score) calculated across ethnic groups for each species, highlighting relative enrichment (red) or depletion (blue) independent of absolute abundance.

To further validate the specific association between *Bifidobacterium* and traditional grain-based staple foods, we also leveraged a longitudinal US cohort where dietary intake was captured using the Automated Self-Administered 24-hour Dietary Assessment Tool (ASA24). We did not find any food items corresponding to idli, thosai, or lontong; nevertheless, our analysis revealed that chapati consumption was associated with significantly higher *Bifidobacterium* abundance compared to non-consumers (*p* < 0.05; **Supplementary Fig. 3b**). These findings mirror our observations in the primary cohort and support the observed diet-microbiome association.

### Cultural Dietary Patterns Among Different Ethnic Groups

The three major ethnic groups demonstrated distinct dietary patterns reflective of their cultural heritage (**Fig. 3b**). Ethnic Indian participants reported higher consumption of traditional whole-grain staple, including chapati (fold change (FC) = 6.559, *q <* 0.001), idli (FC = 5.474, *q <* 0.001) and thosai (FC = 4.81, *q <* 0.001) compared to Chinese and Malay participants. These food items clustered strongly towards the Indian apex in the dietary ternary plot and reflected their strong cultural specificity (**Fig. 4a**, **Supplementary Fig. 4**). Correspondingly, the microbial ternary plot revealed higher relative abundances of *Bifidobacterium*, *Streptococcus*, and *Lactobacillus* species in the ethnic Indians (**Fig. 3c**, **Fig. 4b**), consistent with their metabolic capacity to utilize fiber- and fermentation-derived substrates^58^. Malay participants reported significantly higher intake of noodle dishes and fried foods, including lontong (rice cake; FC = 2.097, *q <* 0.001), begedil (fried potato patties; FC = 3.109, *q <* 0.001), and murtabak (stuffed pancake often consumed during festive times, FC = 4.855, *q <* 0.001). These food items are common staples in the Malay and broader Austronesian cuisine (**Supplementary Fig. 4**)^59^. In contrast, Chinese participants reported higher consumption of animal proteins, including fish (FC = 1.695, *q <* 0.001), seafood (FC = 2.032, *q <* 0.001), and ork (FC = 2.038, *q <* 0.001) (**Supplementary Fig. 4**).

**Figure 4.**
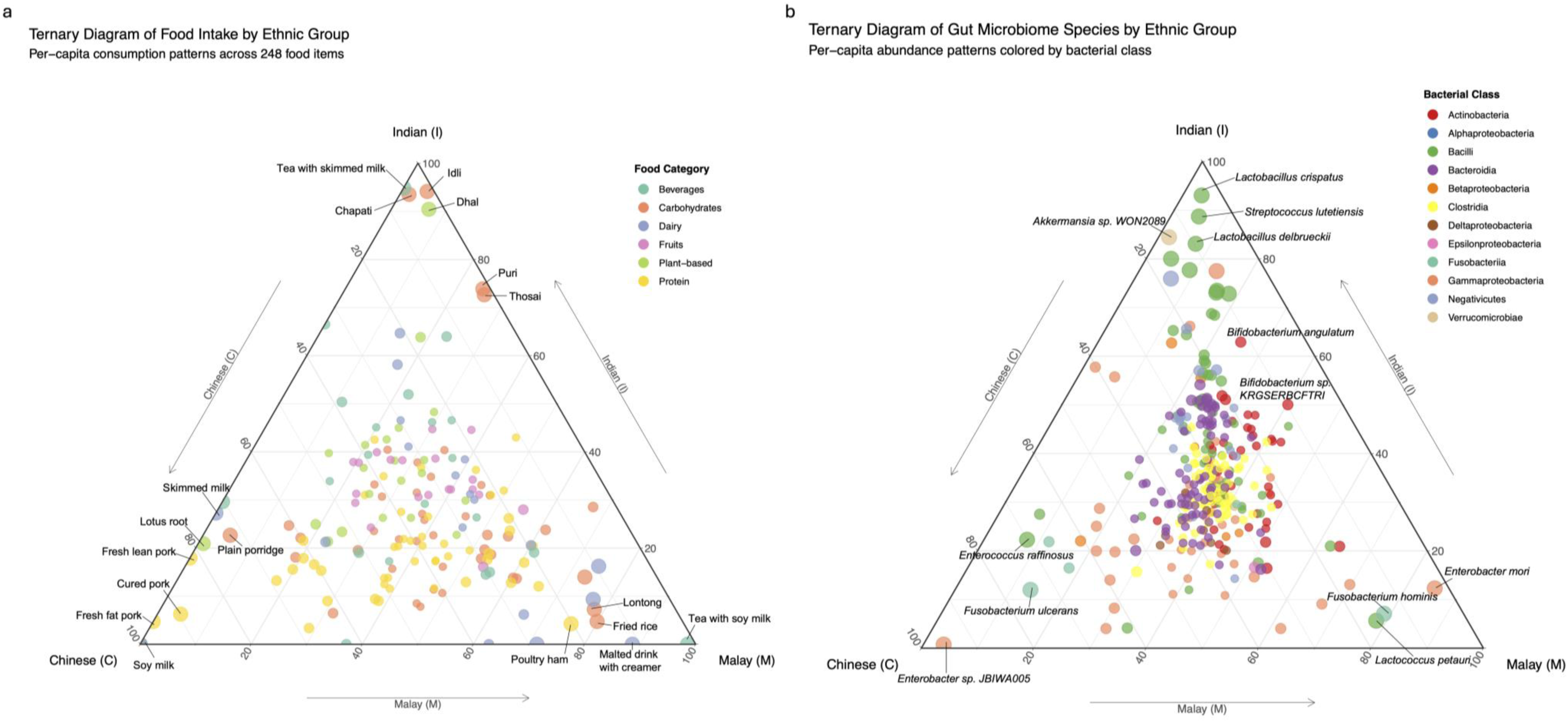
Ethnic variation in diet and gut microbiome composition. (a) Ternary diagram showing per-capita consumption of food items across ethnic Chinese (C), Indian (I), and Malay (M) participants. Points are colored according to food category. (b) Ternary diagram depicting the relative abundance of gut microbial species across the same three ethnic groups, with points colored by bacterial taxonomic class.

### Networks Analysis of Ethnicity-Food-Microbiome Relationships

The correlation networks reveal consistent distinct diet-microbe interactions (**Supplementary Fig. 5**). The network highlights that traditional staple food items, such as chapati (r = 0.232, *q <* 0.05), idli and thosai (r = 0.284, *q <* 0.05) have strong positive correlation with multiple *Bifidobacterium* species as well as the Indian ethnicity (chapati: *r* = 0.437, *q <* 0.05; idli thosai: *r* = 0.465, *q <* 0.05). These staples emerged as central hubs within the network. The ethnicity nodes linked the ethnic Chinese population with higher red meat intake, and the Malay population with their cultural foods such as lontong.

### Stool In-Vitro Microbial Culture Validation

To experimentally validate the observed association between traditional staple foods and *Bifidobacterium*, we employed the INFOGEST *in vitro* digestion and stool *in vitro* culture (SIC) microbial culture model to simulate gastrointestinal digestion and colonic fermentation of thosai, a fermented rice and lentil pancake commonly consumed in the Indian cuisine (**Fig. 5a**). Following the introduction of thosai digesta, we observed a marked expansion of *Bifidobacterium* upon incubation with digested thosai (BRM + thosai P5, P7 and P9), with higher abundance compared with pre-incubation (BRM + thosai P3) and media control (BRM P5, P7 and P9) samples (BH-adjusted *q* < 0.01, **Fig. 5b,c**). Quantitatively, *Bifidobacterium* in the thosai-treated group exhibited a surge up to about 70-fold across different healthy donor microbiome-derived cultures, compared to the comparatively minor and non-significant fluctuations in the control group. A positive monotonic trend was further supported by Spearman correlation (*r* = 0.55, *q* = 0.0015), and linear modelling adjusting for donor effects confirmed passage in the thosai group as a significant predictor of *Bifidobacterium* enrichment (coefficient *β* = 0.215, *q* = 0.00054). This enrichment was consistently observed across all donors despite variable baseline level of the bacteria (**Fig. 5c**). Differential abundance analysis confirmed enrichment of several *Bifidobacterium* species, including *B. longum*, *B. breve*, *B. adolescentis*, and *B. bifidum* with digested thosai, mirroring the species observed in the HELIOS cohort (**Fig. 5d**). These results provide *in vitro* evidence that thosai, a staple of fermented rice-lentil batter, contains substrates that promote *Bifidobacterium* proliferation, thereby validating the possibility of using our diet-microbiome associations to inform targeted dietary interventions.

**Figure 5.**
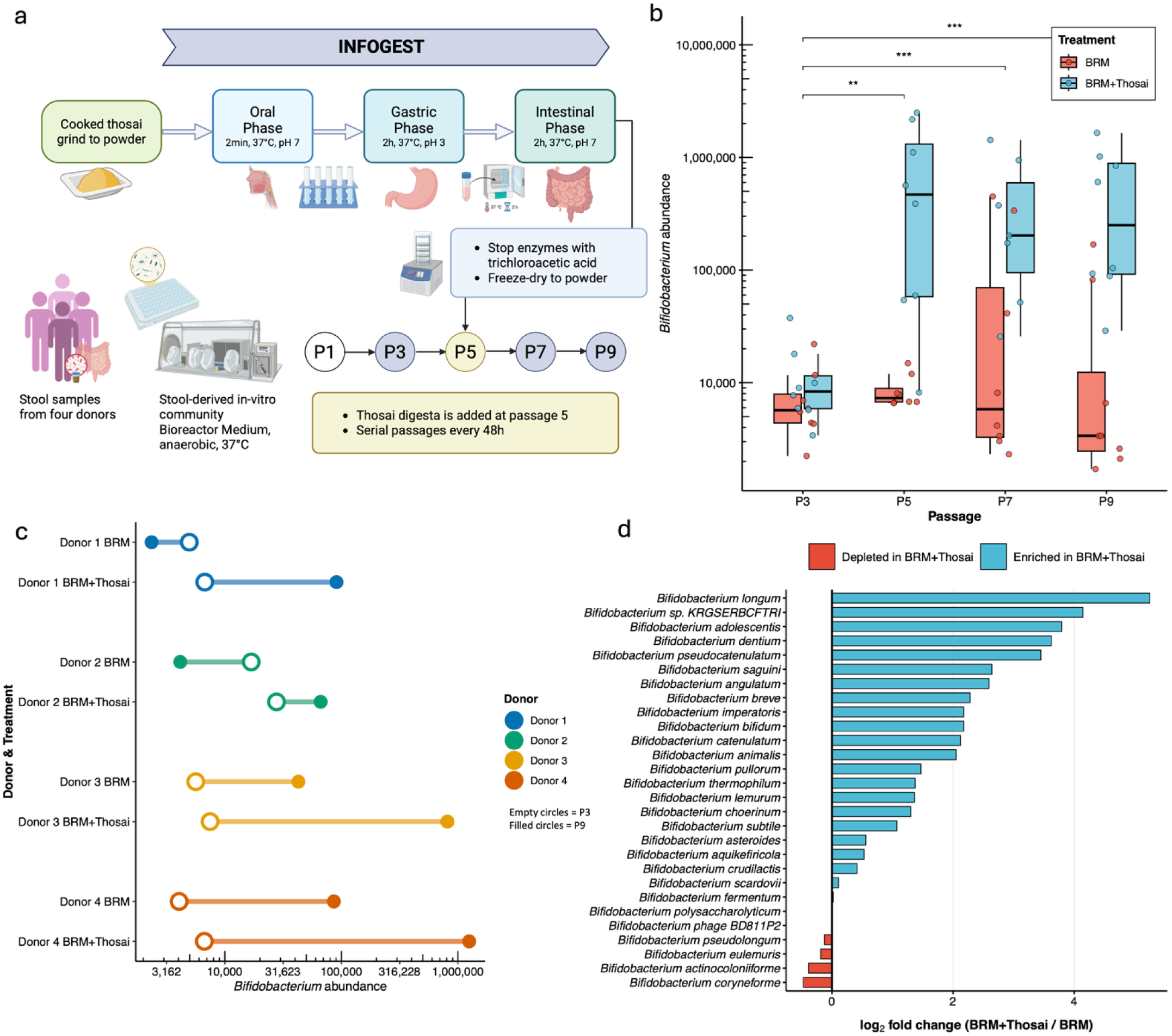
In-vitro experiment results. (a) Schematic overview of the experimental design integrating the INFOGEST digestion model^53^ with subsequent SIC^54^ culture passages. (b) Relative *Bifidobacterium* abundance across SIC passages in BRM control versus cultures supplemented with thosai digesta. Statistical significance between conditions was assessed using Wilcoxon rank-sum tests. (c) Longitudinal changes in *Bifidobacterium* abundance from passages P3 through P9 for each of the four healthy human donors. Empty circles represent P3 and filled circles represent P9. (d) Fold changes of individual *Bifidobacterium* species comparing thosai-treated versus control cultures across donors and passages. Positive values indicate taxa selectively enriched by the thosai digesta, revealing species-specific contributions to the overall *Bifidobacterium* expansion

### Microbiome-derived Disease Risk Scores

To explore the clinical relevance of ethnicity-associated microbiome variations, we explored and calculated microbiome-derived risk scores for five distinct gastrointestinal disorders: CRC, colorectal adenoma, Crohn’s disease, ulcerative colitis, and irritable bowel syndrome based on a previous multi-disease study^46^. Notably, the CRC risk scores revealed a prominent and statistically significant stratification by ethnicity(**Supplementary Figs. 6a-b, Supplementary Table 4**), while the risk scores for other conditions showed non-significant differences between groups.

We observed a clear gradient in the CRC risk scores: ethnic Chinese participants exhibited the highest median risk score, followed by ethnic Malay participants, while ethnic Indian participants scored the lowest (**Supplementary Fig. 6c**). To ensure the robustness of this finding, we applied a second, independent CRC-specific scoring framework derived from a large-scale meta-analysis^47^. This validation step corroborated our initial results, reproducing the same hierarchical trend among the three ethnic groups (**Supplementary Fig. 6d**). Correspondingly, this microbiome-inferred risk stratification mirrors the epidemiological landscape in Singapore, with a concordance between the microbiome-derived risk scores and national health statistics reporting the highest CRC incidence in the ethnic Chinese and the lowest incidence among ethnic Indian populations^60^.

## Discussion

This study provides a comprehensive exploration of the intricate relationship between diet, ethnicity, and gut microbiome in a multi-ethnic urban population in Singapore. By leveraging data from the HELIOS cohort, this research captures the dietary and lifestyle diversity of three major ethnic groups – Chinese, Indian, and Malay – while examining their microbiome within a shared urban geography. Rather than comparing physically distant populations across different countries, our study minimizes geographic and infrastructural confounding to delineate the relationship between cultural dietary habits and gut microbiome. Importantly, these ethnic groups are relevant to the Austronesians, South Indians, and Han Chinese populations that represent over 2.5 billion people, comprising nearly one-third of the world’s population and underscoring the potential far-reaching implications of these findings. Our work establishes a reference framework for microbiome research in the Asia-Pacific region, which remains underrepresented compared to Western cohorts.

Our findings revealed distinct yet overlapping microbiome profiles across the three ethnic groups^61,62^. In our cohort, all participants resided within the same highly urbanized city in Singapore, characterized by shared built infrastructure, centralized water and sanitation systems, broadly similar food supply chains, and common healthcare access. Previous studies have shown that many of these lifestyle factors, including urbanization^63^, geography^64^, built-in environment and water source^65^ are most influential in shaping gut microbial communities. Within this context, these common environmental features may represent a plausible backdrop for the considerable overlap observed in microbial composition^66^. However, each ethnic group retained unique microbial features that paralleled their dietary practices. This microbial distinctiveness, despite decades of co-residence in the city-state, highlights the resilience of culture-specific dietary signatures in shaping gut ecology.

We observed that the ethnic Indian population has a higher intake of grain-based staple foods like idli and thosai, which correlate with a higher abundance of *Bifidobacterium* in their gut microbiome. This bacterial enrichment in the Indian population was consistently observed in two independent Malaysian cohorts^26^, which share similar geography and constituent ethnic groups. This relationship may reflect the metabolic capacity of bifidobacteria to utilize lactate^67,68^. In the intestines, lactic acid bacteria and host epithelial cells produce lactate, which bifidobacteria can metabolize through the lactate dehydrogenase (LDH) pathway to convert into pyruvate and subsequently acetate^68,69^. Beyond lactate, thosai also contains resistant starches and exopolysaccharides, which may further contribute to their growth as efficient fiber consumers^70,71^. Together with the American cohort results and our *in vitro* digestion and microbial culture models, these findings suggest that these staple foods may promote gut microbial shifts through both lactate metabolism and fiber fermentation.

Nevertheless, while *Bifidobacterium* is often regarded as a probiotic, its presence does not necessarily equate to better metabolic health. For instance, ethnic Indians in Singapore experience a higher prevalence of diabetes compared to Malays and Chinese^72^. These findings suggest that the effects of *Bifidobacterium* may be modulated by other lifestyle and host factors. Indeed, prior studies have shown that Indians consumed fewer fruits and vegetables overall^73^, which may attenuate the potential benefits of *Bifidobacterium*. These findings emphasize that microbial shifts cannot be interpreted in isolation but must be considered within broader lifestyle habits and host susceptibility.

The gut microbiome is now recognized as a causal and modulatory factor in CRC, with distinct bacterial taxa experimentally validated to promote tumorigenesis or exert protective effects^74,8^. Our risk-score analysis revealed higher microbiome-derived CRC scores in ethnic Chinese than those of Malay and Indian subjects. This parallels the population-level incidence rates in Singapore, with ethnic Chinese having the highest and Indian having the lowest CRC incidence^75^. Notably, Indian participants displayed marked enrichment of *Bifidobacterium* and *Akkermansia*, both regarded as beneficial commensals with putative anti-tumor and muco-protective effects^76,77^. Such a microbial configuration with probiotic-associated taxa may help establish a milieu that reduces CRC risk. Collectively, these observations highlight the complex interplay between habitual diet and gut microbial composition which could track closely with population-level differences in disease risks.

This study depicts the microbiome in several Asian ethnic groups and gives insights into precision nutrition strategies, considering ethnicity, dietary habits, and microbiome composition. Clearly, a single-focus approach addressing one genetic or environmental factor is inadequate, as it fails to capture the complex interplay between these factors and their interacting impact on health. While diet is a major determinant of microbiome composition, the microbiome itself mediates how nutrients are metabolized to influence health outcomes. Similarly, cultural dietary preferences, genetic predisposition, and lifestyle behaviors all interact to shape microbiome diversity and functionality. An integrative framework that recognizes these multi-layered interactions is essential for addressing ethnicity-specific health disparities and designing effective public health interventions. Our findings provide the foundation for designing culturally tailored, population-specific dietary interventions, where traditional food practices can be leveraged or modified to optimize microbial profiles and health. By embedding microbiome science within the broader context of public health and cultural diversity, these insights have the potential to inform strategies not only for Singapore, but also for the wider populations of different ethnic groups across Asia and beyond.

## Conclusion

Our study highlights diet-microbiome relationships as a key factor in health and disease, emphasizing the need to consider diet and microbiome as an integral system. Using a unique national cohort in a city-state, our findings revealed distinct relationships between ethnic groups, culture-relevant diets, and specific microbial taxa. By incorporating these multi-dimensional data, we provide evidence that cultural context and microbial ecology should be considered in developing precision nutrition strategies, which may help improve health outcomes and address health disparities This study is relevant for Asia Pacific region where dietary practices and disease patterns differ substantially from Western populations. Future research should move beyond cross-sectional designs and include longitudinal cohorts outside an urbanized setting, to capture diet-microbiome interactions that evolve across time, geography and socioeconomic transitions. This study provides a foundation for culturally informed, population-specific strategies that harness microbiome science to advance precision public health and guide global nutrition and health policies.

## Supporting information

The author list of the HELIOS Study Team

## Data Availability

All data produced in the present study are available upon reasonable request to the authors. The HELIOS study individual-level datasets are not publicly available due to data privacy regulations. Researchers may apply for access through HELIOS Data Access Committee. Summary-level datasets are available as supplementary tables including microbiome differential abundance analysis results, food-microbes partial correlation coefficients, and disease risk scores.

## Author contributions

RZ: Investigation; Formal analysis; Data curation; Writing – original draft. XS, THM, KRA, DYL, JXK, JJYT, SHCH: Formal analysis; Data interpretation. BCCL, JJYS, CWC, NN, YL, JCC: Resources; Data curation; Infrastructure support. NN, YL, JCC, SHW: Conceptualization; Methodology; Formal analysis; Data interpretation; Writing – review & editing. HELIOS Study Team: Resources; Data curation; Project administration. All authors contributed intellectual input, critically reviewed the manuscript, and approved the final version.

## Acknowledgement

We thank the participants of the HELIOS study and the HELIOS operation team for recruitment, organization and data/sample collection.

The HELIOS study (NTU IRB: 2016-11-030) is supported by Singapore Ministry of Health’s (MOH) National Medical Research Council (NMRC) under its OF-LCG funding scheme (MOH-000271 and MOH-001792) and NMRC through National Cohorts Office (P2022-02-03) and the National Precision Medicine (NPM) Programme. NPM Programme Phase II is supported by the National Research Foundation, Singapore (NRF) under the RIE2020 White Space (MOH-000588 and MOH-001264) and administered by the Singapore Ministry of Health through the National Medical Research Council (NMRC) Office, MOH Holdings Pte Ltd. NPM Programme Phase III is supported by the Singapore Ministry of Health through the NMRC Office, MOH Holdings Pte Ltd under the NMRC RIE2025 NPM Phase III Funding Initiative (MOH-001734). HELIOS is also supported by intramural funding from Nanyang Technological University, Lee Kong Chian School of Medicine and National Healthcare Group.

The other parts of the study are supported by grant NMRC OF-LCG (MOH-001326), NMRC CS-IRG (MOH-001353), NMRC OF-YIRC (MOH-001732), Centre for Microbiome Medicine, Wang Lee Wah Memorial Fund, Ministry of Education Singapore (MOE-T2EP30221-0003, 2019-T1-001-059, RG30/23), the National Research Foundation (NRF2020-THE003-0006), the Future Ready Food Safety Hub of NTU Singapore, and Asian Institute of Modern Agrinomy (AIMA), LKCMedicine Healthcare Research Fund (Diabetes Research), established through the generous support of alumni of Nanyang Technological University, Singapore.

## Code availability

Data processing, analysis and visualization were performed using R (v4.5.0). Codes for analyses and visualization are available via GitHub at https://github.com/jaspershen-lab/HELIOS-project

---

**Supplementary Figure 1.**
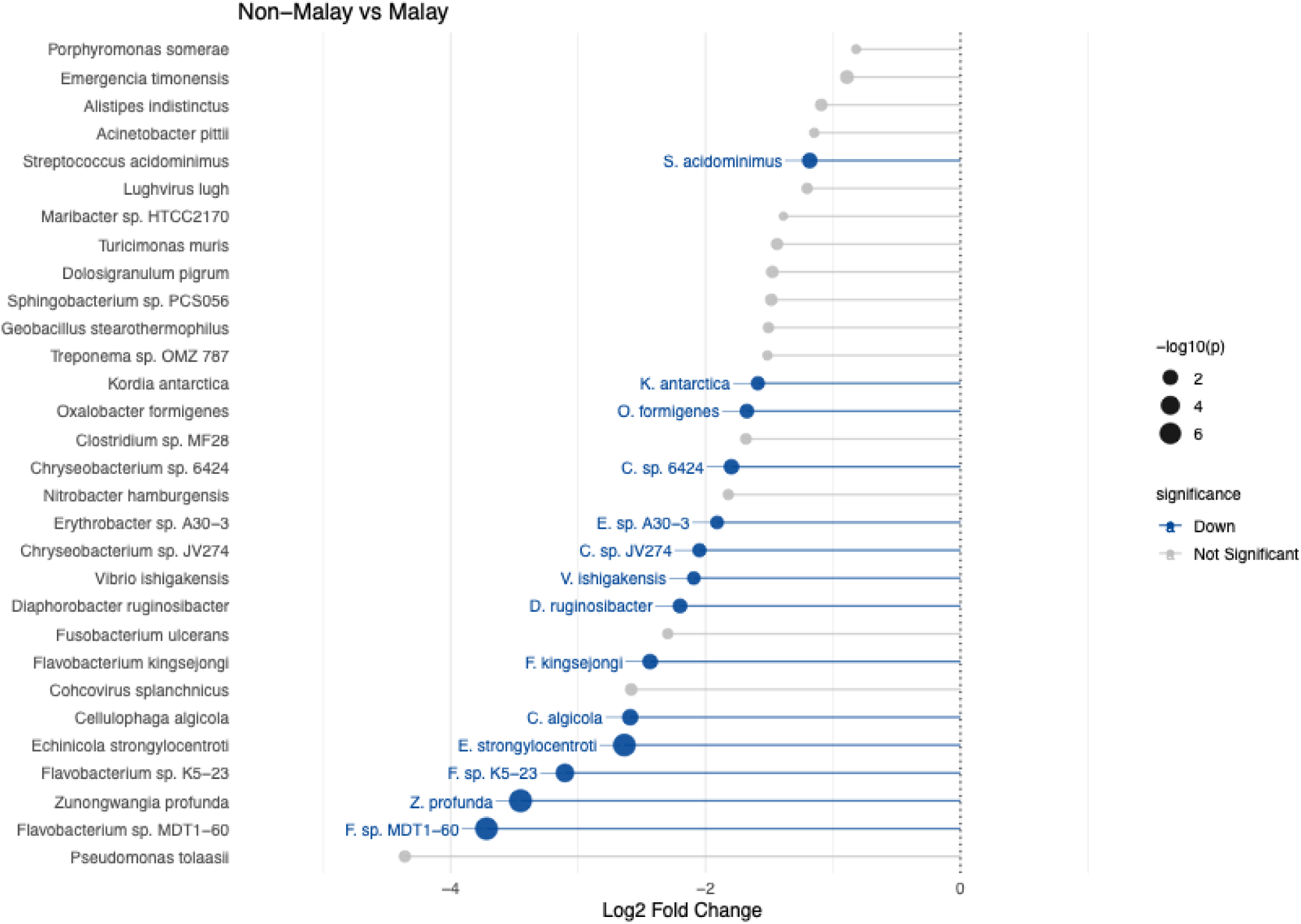
Differential abundance analysis of microbiome taxa comparing Malay and non-Malay individuals, visualized as a lollipop plot. Each point represents a microbial species, with the x-axis indicating the effect size (log₂ fold change). Blue points and lines represent species depleted in Malay individuals significantly (p value < 0.05). Point size corresponds to the relative abundance of each species in the dataset. Species are ordered vertically according to effect size, with labels provided for the most strongly enriched taxa.

**Supplementary Figure 2.**
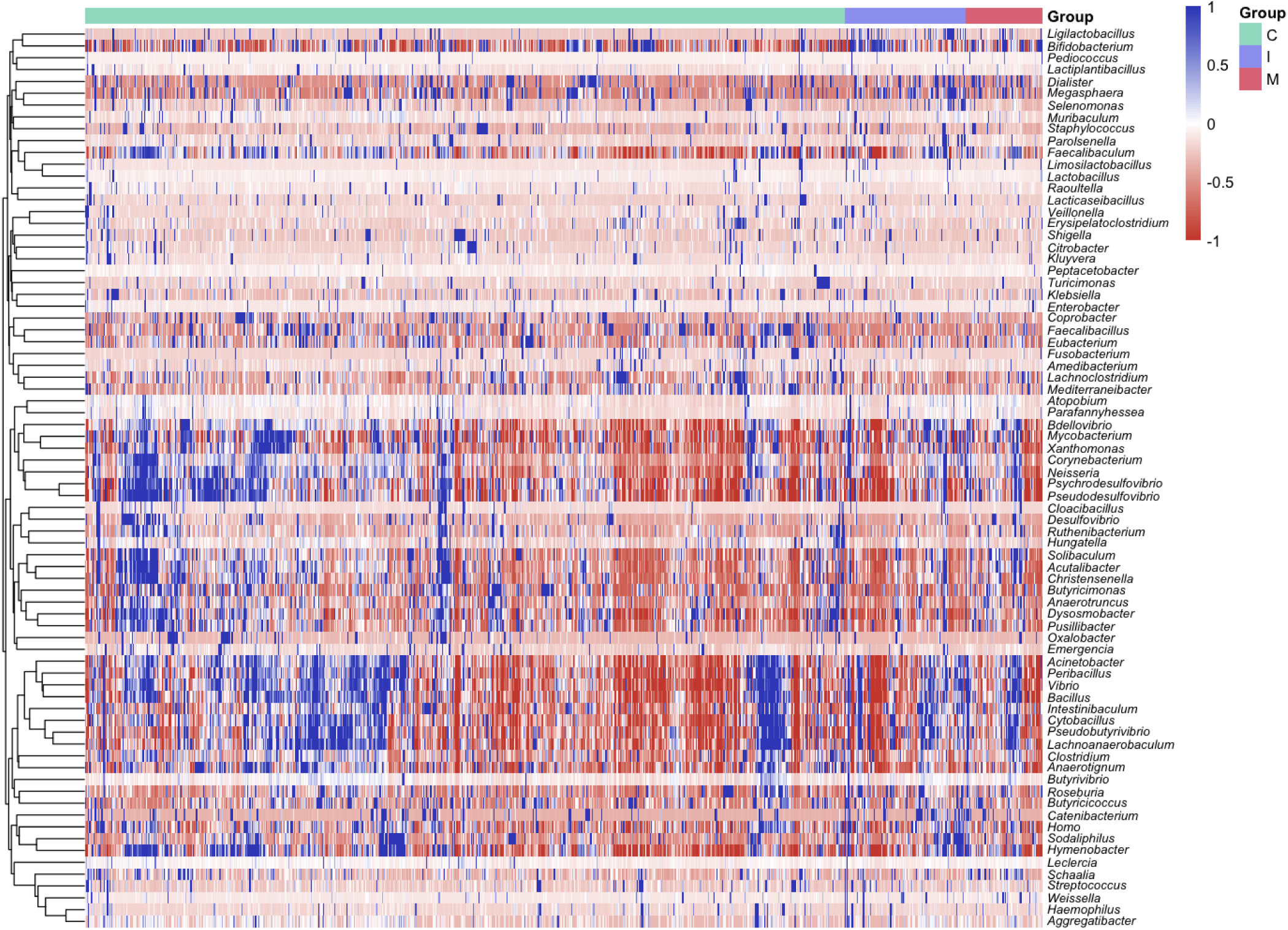
Genus-level heatmap of microbiota abundance differences across ethnic groups in the HELIOS cohort. This heatmap displays the scaled relative abundances (z-scores) of genera that were identified as significantly differentially abundant across the ethnic groups based on differential abundance analysis. Rows represent genera and columns represent individual participants, ordered by hierarchical clustering using Bray-Curtis distance. Colors indicate standardized abundance values, with red representing higher-than-average abundance and blue representing lower-than-average abundance within each genus.

**Supplementary Figure 3.**
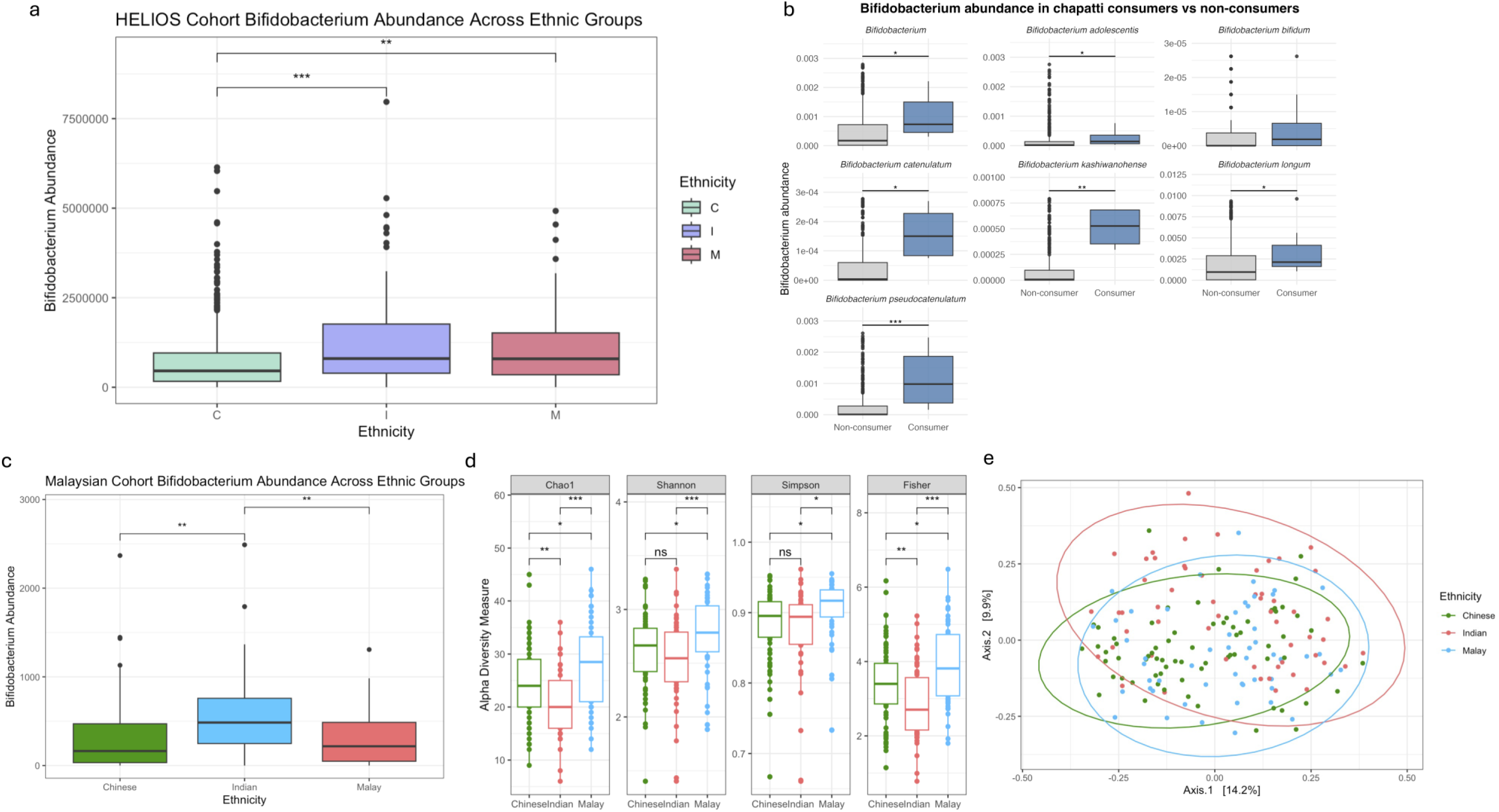
(a) *Bifidobacterium* abundance across Chinese, Indian, and Malay participants in the HELIOS cohort. (b) In ASA24 cohort, Comparison of Bifidobacterium abundance between chapatti consumers and non-consumers. (c) Validation of ethnic differences in *Bifidobacterium* abundance in an independent Malaysian cohort. (d) In Malaysian cohort, comparison of alpha-diversity metrics (Chao1 richness, Shannon diversity, Simpson diversity, and Fisher index) among Chinese, Indian, and Malay participants. (e) In Malaysian cohort, principal coordinates analysis (PCoA) based on Bray–Curtis dissimilarity illustrating ethnic differences in overall gut microbial community structure.

**Supplementary Figure 4.**
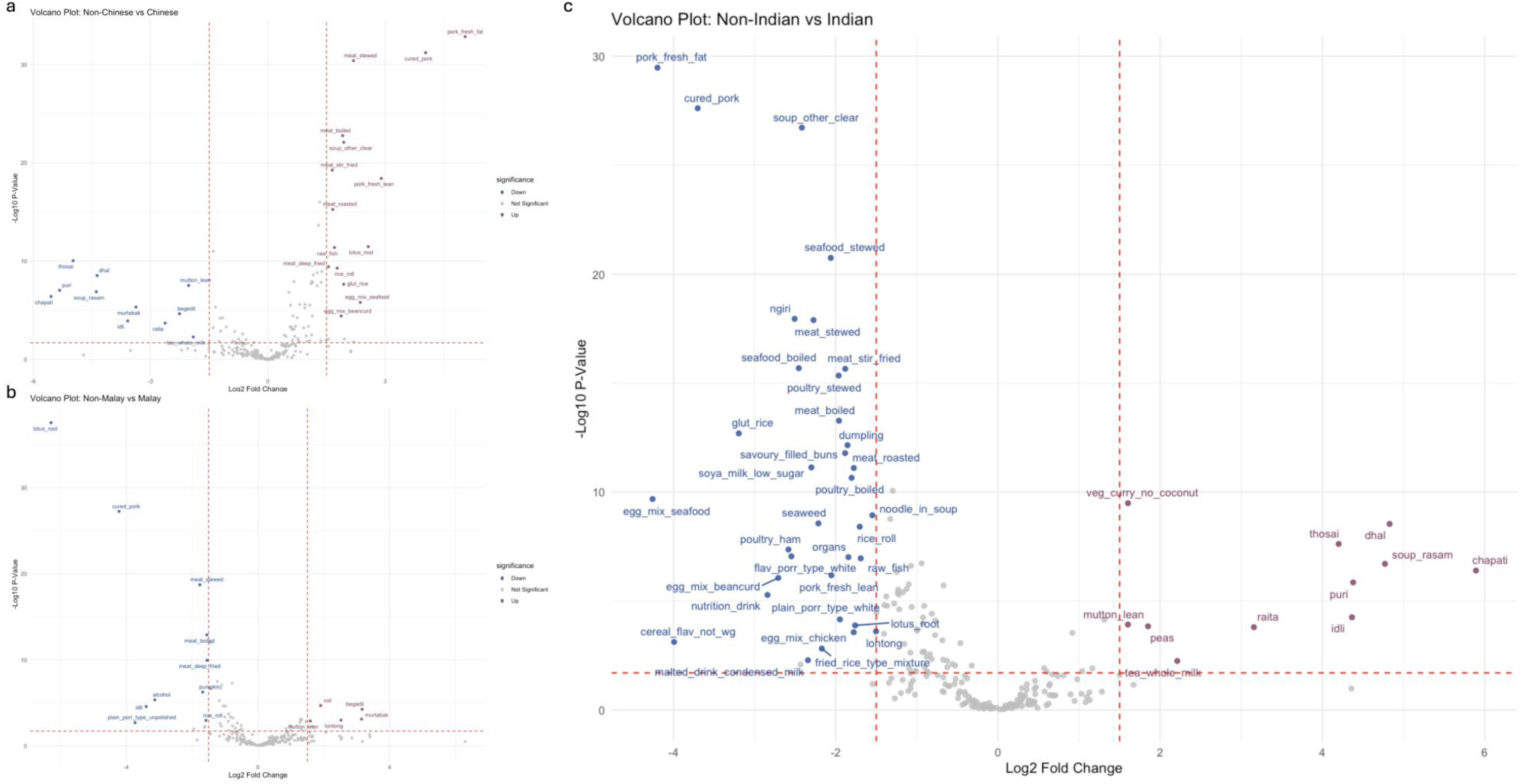
Differential food consumption patterns among ethnic groups. Volcano plots illustrating differences in food consumption between (a) Chinese and non-Chinese, (b) Malay and non-Malay, and (c) Indian and non-Indian groups. Statistically significant differences in dietary intake between groups are highlighted, with key food items exhibiting differential consumption patterns.

**Supplementary Figure 5.**
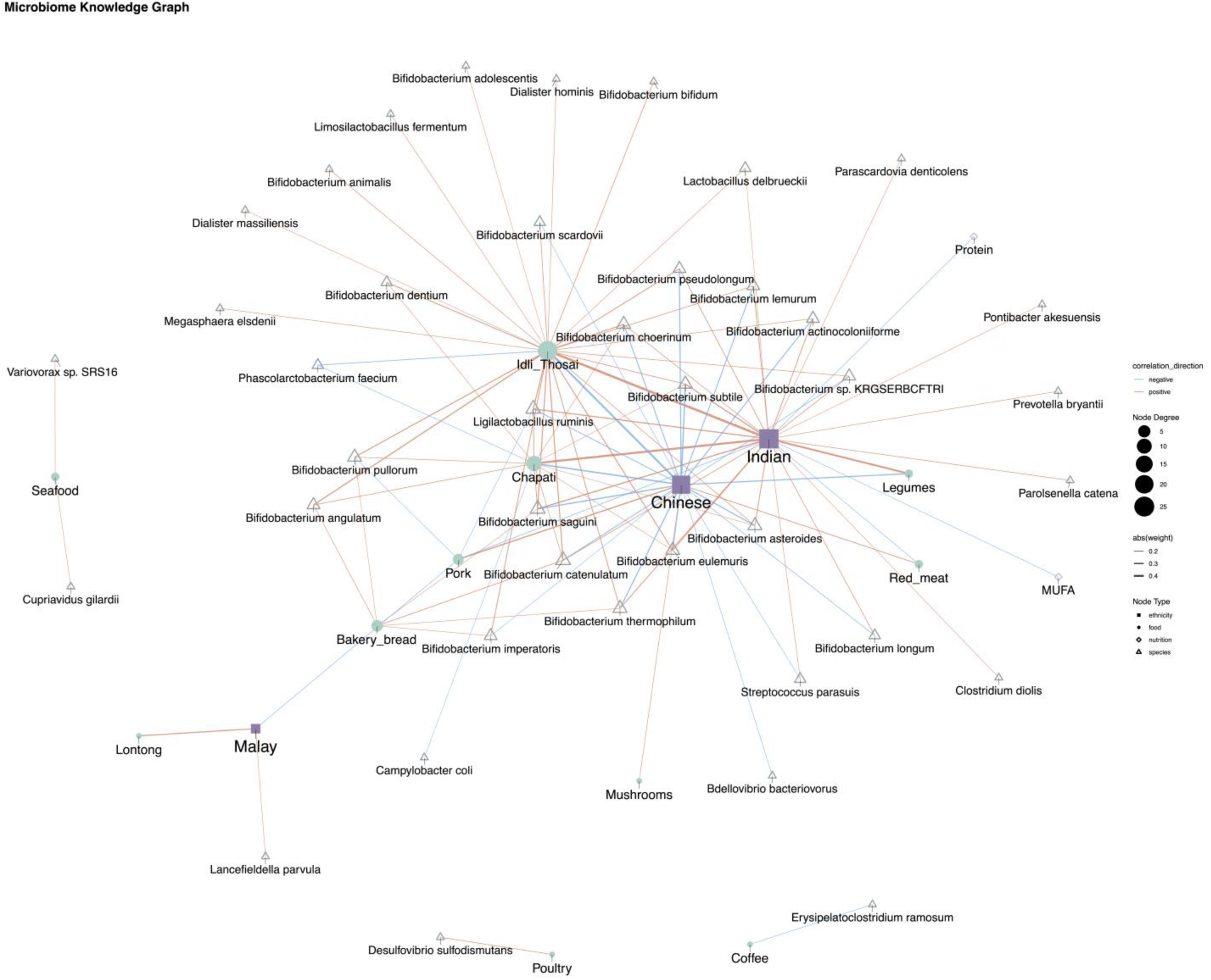
Correlation networks between dietary intake, ethnicity, and gut microbial species. Network visualization of associations between food intake, nutrient consumption, and microbial species. Relationships involving ethnicity (encoded as dummy variables) are assessed using Point-Biserial correlation, while all other associations are evaluated using Spearman’s correlation. Edges represent statistically significant correlations (p<0.05 and |correlation coefficient| > 0.13, adjusted for multiple comparisons), with red edges indicating positive correlations and blue edges indicating negative correlations. The thickness of each edge reflects the strength of the absolute correlation. This network highlights distinct patterns of food-microbe and nutrient-microbe interactions across ethnic groups.

**Supplementary Figure 6.**
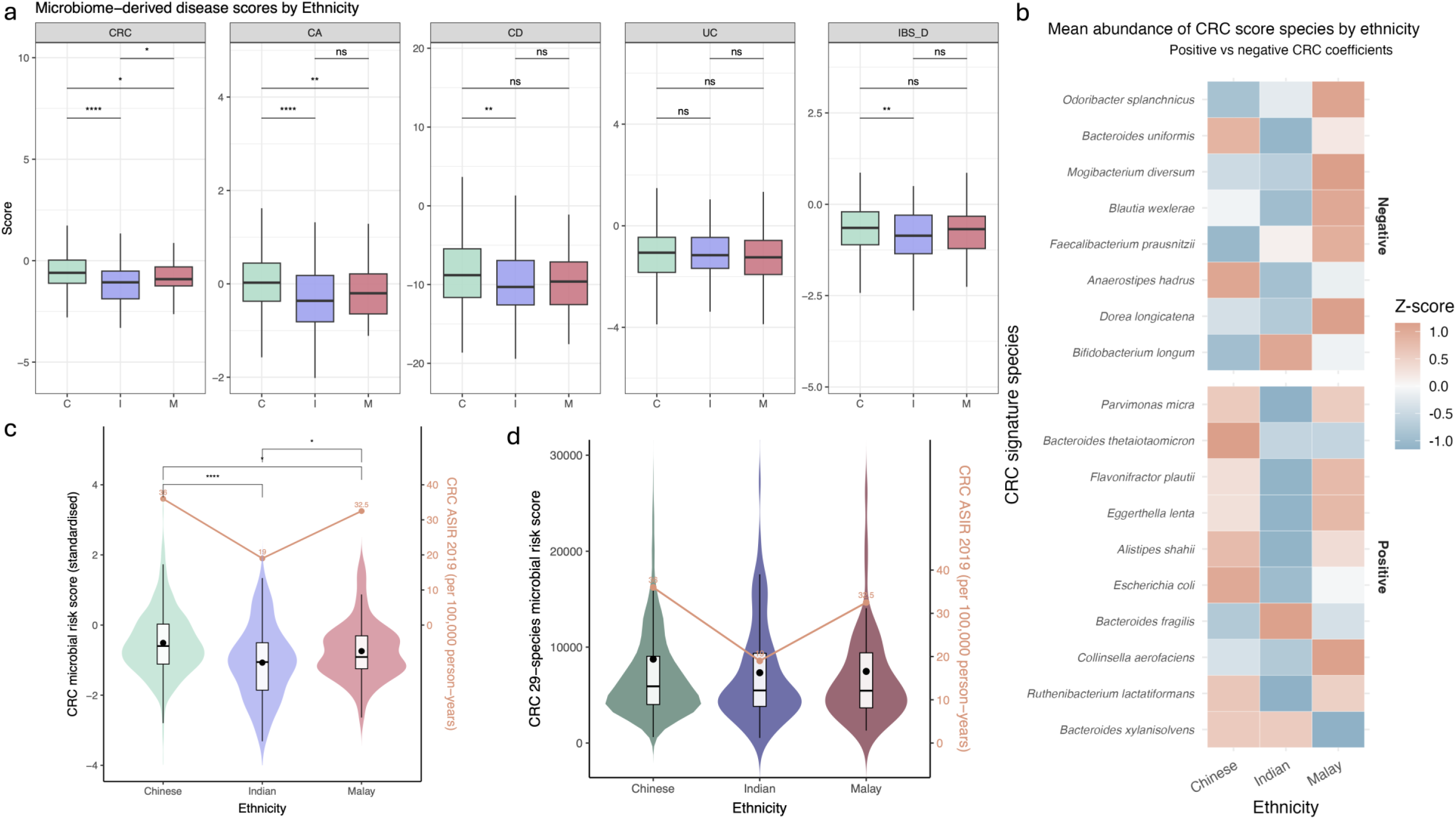
Ethnic variation in microbiome-derived colorectal neoplasia scores. (a) Boxplots display the distribution of microbiome-derived risk scores for five gastrointestinal disorders—colorectal cancer (CRC), colorectal adenoma (CA), Crohn’s disease (CD), ulcerative colitis (UC), and irritable bowel syndrome (IBS-D), across ethnic populations. (b) Heatmap of mean-centred z-scores for cancer-associated microbial species across ethnic groups. Species are grouped according to whether they carry positive or negative coefficients in the original colorectal neoplasia models. Orange shades indicate relative enrichment within an ethnic group, whereas blue shades indicate relative depletion. (c) Violin and boxplots showing distribution of the 50-species microbiome-derived colorectal neoplasia score across ethnic Chinese, Indian, and Malay participants^38^. Orange horizontal lines denote ethnic group means. (d) Distribution of the 29-species colorectal neoplasia score stratified by ethnic groups^3^

